# Integrating Isoniazid Preventive Therapy into the Fast-Track HIV Treatment Model in Urban Zambia: A Proof-of -Concept Pilot Project

**DOI:** 10.1101/2022.07.23.22277950

**Authors:** Mpande Mukumbwa-Mwenechanya, Muhau Mubiana, Paul Somwe, Khozya Zyambo, Maureen Simwenda, Nancy Zongwe, Estella Kalunkumya, Linah Kampilimba Mwango, Miriam Rabkin, Felton Mpesela, Fred Chungu, Felix Mwanza, Peter Preko, Carolyn Bolton-Moore, Samuel Bosomprah, Anjali Sharma, Khunga Morton, Prisca Kasonde, Lloyd Mulenga, Patrick Lingu, Priscilla Lumano Mulenga

## Abstract

**Introduction:** Most people living with HIV (PLHIV) who are established on treatment in Zambia receive multi-month prescribing and dispensing (MMSD) of antiretroviral therapy (ART) and are enrolled in less-intensive differentiated service delivery (DSD) models, such as Fast Track (FT), in which clients pick up ART every 3-6 months and make clinical visits to health facilities every 6 months. In 2019, Zambia introduced Isoniazid Preventive Therapy (IPT) with scheduled visits at 2 weeks and 1, 3, and 6 months. The asynchronous IPT and HIV appointment schedules were inconvenient and not client centered. In response, we piloted integrated MMSD/IPT in the FT HIV treatment model.

**Methods:** We implemented and evaluated a proof-of-concept pilot project at one purposively selected high-volume facility in Lusaka, Zambia between July 2019 and May 2020. We sensitized stakeholders, adapted training materials and standard operating procedures, and screened adults in FT for TB as per national guidelines. Participants received structured TB/IPT education, a 6-month supply of isoniazid and ART, an aligned 6^th^ month IPT/MMSD clinic appointment, and phone appointments at 2 weeks and months 1-5 following IPT initiation. We used descriptive statistics to characterize IPT completion rates, phone appointment keeping, side effect frequency and Fisher’s exact test to determine if these varied by participant characteristics. Notes from monthly meetings and discussions were used to synthesize key lessons learned.

**Results:** 1,167 clients were screened for eligibility and 818 (70.1%) were enrolled. Two thirds (66%) were female, median age was 42 years, and 56.6% had been on ART for ≥ 5 years. 738 (90.2%) completed a 6-month course of IPT and 66 (8.1%) reported IPT-related side effects. 539 clients (65.9%) attended all 7 telephone appointments. There were no significant differences in these outcomes by age, sex, or time on ART. Lessons learnt include the value of promoting project ownership, client empowerment, securing supply chain, adapting existing processes, and cultivating a collaborative structured learning environment.

**Conclusions:** Integrating multi-month dispensing and telephone follow up of IPT into the FT HIV treatment model is a promising approach to scaling-up TB preventive treatment among PLHIV, although limited by barriers to consistent phone access.

## Introduction

Globally, tuberculosis (TB) is the direct cause of one-third of all human immunodeficiency virus (HIV) related deaths and the leading cause of death among people living with HIV (PLHIV). ^1^ PLHIV account for approximately 10% of the 10 million annual new TB cases and on average, have about 21-fold (CI: 16–27) higher risk of developing TB than HIV-negative persons.^2-5^ Zambia has one of the highest TB burdens in sub-Saharan Africa (SSA) and is ranked among the top 30 TB control priority countries by the World Health Organization (WHO).^1^ In 2021, the estimated 59,000 new cases of TB in Zambia contributed to approximately 14,800 deaths among Zambians, of which 9,100 (62%) occurred among PLHIV.^2,3^

The WHO Global End TB strategy recommends provision of TB preventive treatment (TPT) for PLHIV with the aim of eliminating TB by 2030. Whilst a priority and cost-effective, TPT coverage and uptake has been low among PLHIV^3^ and current TPT coverage remains well under WHO’s 2025 coverage target (≥ 90%). ^6-10^ Commonly cited barriers for low uptake and coverage include stock-outs, perceived fears of poor adherence and isoniazid (INH) resistance, inability to prevent and monitor adverse events, a lack of local or national commitment, and perceived difficulty of ruling out active TB. ^11-13^

In 2019, the Zambian Ministry of Health (MoH) adopted the WHO-recommended TPT strategy.^2^ The MoH TPT guidance provided for Isoniazid Preventive Therapy (IPT) dispensed at 2 weeks and 1, 3 and 5 months for PLHIV established on treatment, defined as non-pregnant adults on ART for greater than 6 months, with VL less than 1000 copies/mL, and without opportunistic infections or active TB. This same population was eligible to enroll in Differentiated Service Delivery (DSD)^14^ for HIV treatment, enabled by multi-month (3-6 months) scripting and dispensation (MMSD) of ART, shown to improve retention and viral suppression.^15^

One of the DSD models utilizing MMSD in Zambia is the Fast-Track (FT) model, an individual facility-based model in which clients visit health facilities every three months, alternating expedited medication pick up visits and clinical visits (Figure 1).^14^ As of June 2021, 13% of people on ART in Zambia had enrolled in FT.^15^ Although integration of IPT into this model is an important part of TPT scale up, the asynchronous appointment schedules for ART and IPT threatened DSD’s “client centered” approach and retention in HIV-TB care.

**Figure 1:**
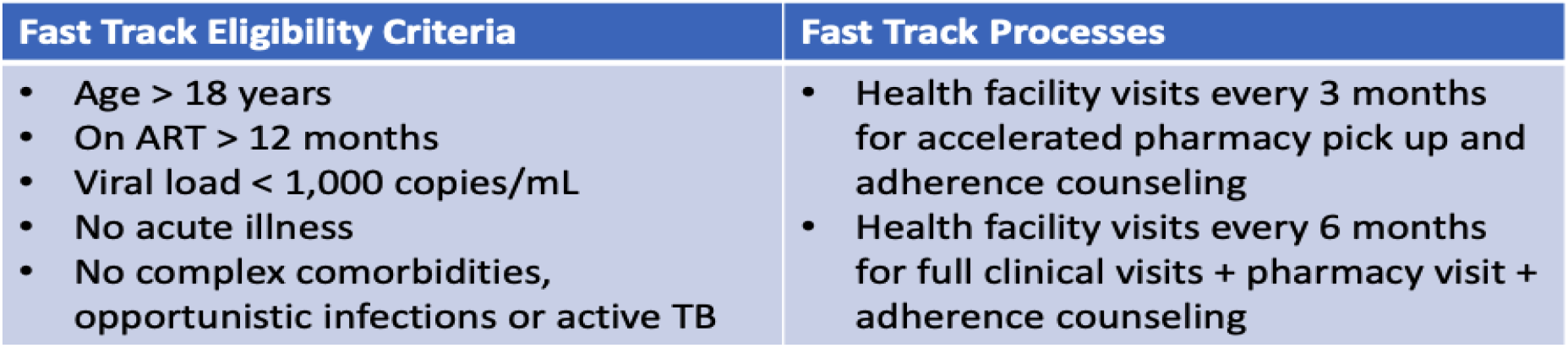
The Fast -Track HIV Treatment Mode.

Integrating multi-month dispensing of IPT into a rapidly growing DSD model for HIV care offers a promising public health strategy to improve TPT uptake and prevention of TB among PLHIV.^16^ DSD models with longer refill intervals have demonstrated high rates of long-term retention where 90% remained in care at four years compared to less than 70% in traditional facility-based HIV programs with monthly visits. ^17-20^

While case findings from SSA countries have shown that integration of MMSD TPT into DSD models is feasible, the IPT refills were limited to a maximum of three months with minimal evidence of adherence support and side effect monitoring at shorter intervals via phone. ^14,21^ Furthermore, the use of mobile phones for continuous PLHIV monitoring and adherence support and provision of 6MMSD TPT is silent in framework documents for programmatic considerations of TPT implementation in DSD models.^21^ In response, we designed a proof-of-concept pilot project that integrated MMSD and IPT services in the FT DSD model by synchronizing long refills of IPT (6 months MMSD) and visit schedules and utilized phone calls to provide adherence support and side effect monitoring. The approach was piloted at a single health facility, given limited resources and the desire to start small and learn from experience.

Objectives included documenting the inputs, processes and enabling factors and challenges involved in launching this new approach to TPT delivery, as well as client uptake, completion of phone check-ins, and self-reported TPT completion rates and side effects. We did not attempt to compare TPT outcomes *vs*. standard of care or to assess generalizability.

## Methods

### Study site

We purposively selected a high-volume HIV clinic serving more than 10,000 PLHIV active on ART (including 5,000 enrolled in FT) located within an urban primary level health center in Lusaka. We believed this site to be relatively common in our country (even if not typical) to quickly learn and generate valuable lessons that could contribute to the dialogue about integration of TPT into DSD models.

### Study design and participants

The pilot was implemented in three phases: preparatory, enrollment, and intervention phase (Figure 2). Inclusion criteria included age over 18 years, being established on ART as per national guidelines, enrolled in FT, having access to a functioning phone, and willing to participate. Exclusion criteria included recent or current IPT use, recent or current TB treatment, current TB symptoms, and pregnancy. All clients that came to the facility during the one-month enrollment period (September 2019 – October 2019) were eligible to enroll.

**Figure 2:**
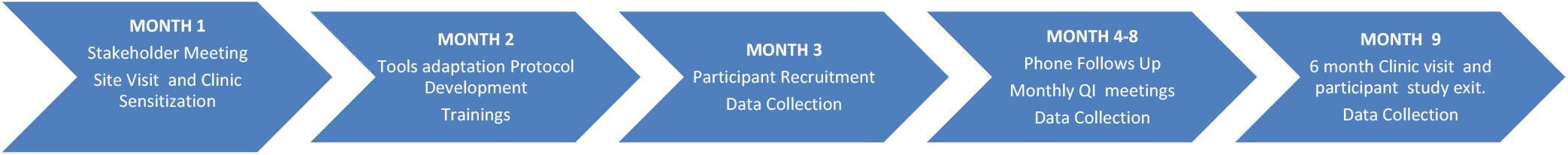
Project Implementation Timeline.

We piloted the intervention, consisting of an adjusted IPT visit schedule; baseline education and counselling; dispensing a 6 month-supply of both IPT and ART; phone check-ins at 2 weeks and then monthly for 5 months; and a synchronized in-person follow-up visit for HIV and IPT care at 6 months (Figure 3).

**Figure 3:**
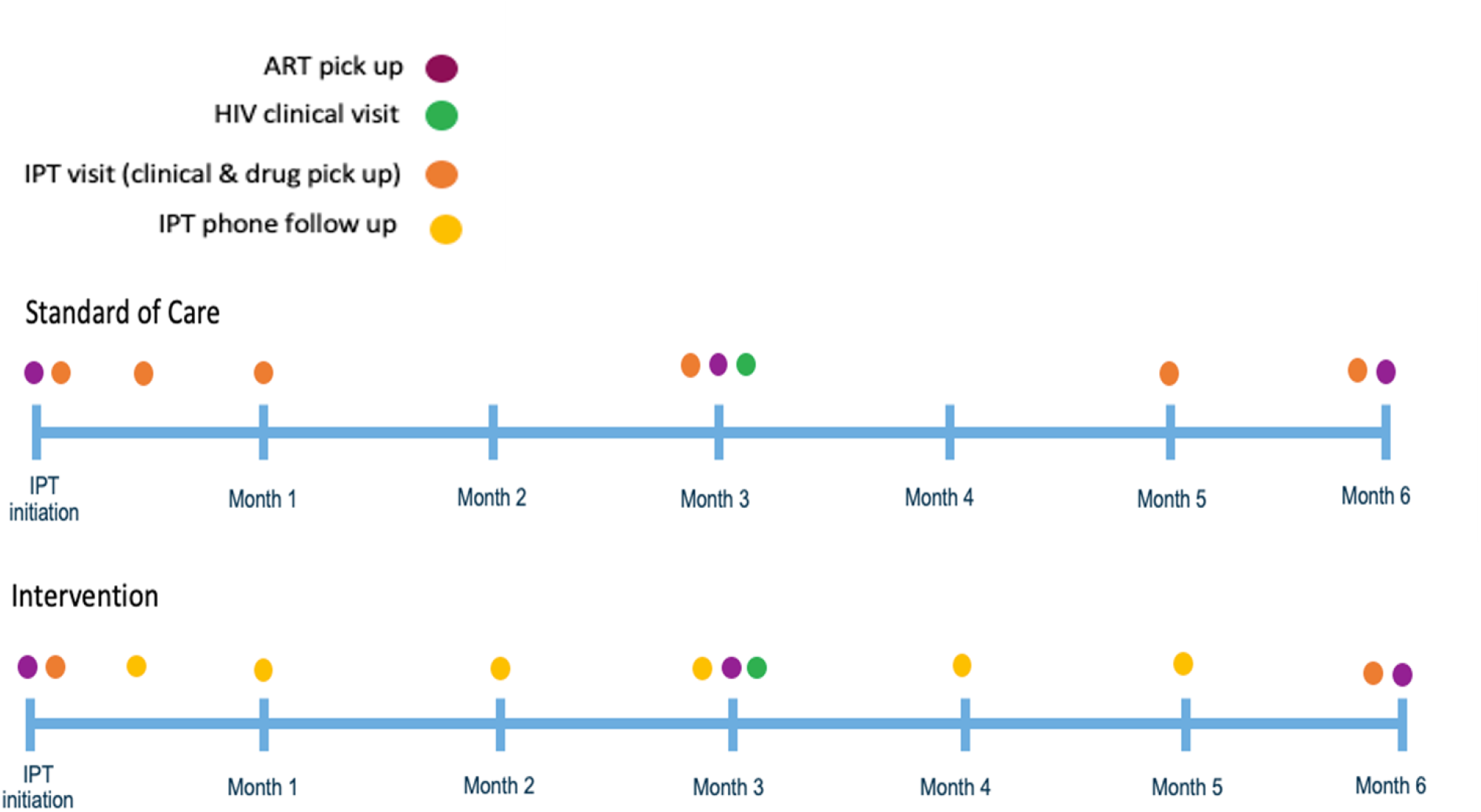
Standard of Care vs. Intervention.

### Study procedures

Before protocol development, the pilot team carefully documented FT model processes and reviewed the existing supply chain and infrastructure. This information was used to design the intervention model (Figure 3) and to identify the appropriate facility staff to provide screening, counselling, enrolment, IPT initiation and follow-up services.

Cognizant that a strong supply chain system that ensures availability of both INH and Pyridoxine (Vitamin B6) was pivotal for project success,^22^ the project team engaged the NTP Pharmacist and health facility Pharmacist to secure a constant supply of commodities. The project team developed, and adapted training materials, standard operating procedures (SOPs) and data collection tools and conducted stakeholder sensitization with Civil Society Organizations (CSOs), healthcare workers (HCWs) and PLHIV to ensure initial and ongoing success of IPT incorporation into FT. Conducted in-person didactic training reorientated health centre staff to standardized screening for IPT eligibility and management of patients on IPT including symptom screening and counselling. In addition, community health workers (CHWs) were trained on conducting structured phone follow-ups and provide referral for clinical consultations.

During the month-long enrolment phase (September 2019 - October 2019), HCWs routinely screened adults in FT for pilot project eligibility. Eligible clients were invited to enroll in the integrated IPT/ART model; participants then received structured health education messages on the importance and benefits of IPT as well as identification, reporting and management of TB symptoms/expected side effects. Enrollees received 6 months dispensation of INH (300mg), pyridoxine (Vitamin B6) and their current ART regimen, and an appointment to return in 6 months.

During the intervention phase, CHW conducted phone follow-ups at 2 weeks, 4 weeks, and then once monthly for 5 months. Structured checklist and script was utilized to assess IPT adherence and any related side-effects and concerns/experiences (see Additional file, Section one). When participants were not available by phone after two failed attempts on two separate days, the CHWs followed up with physical visits to their homes using the address provided by the clients at enrollment. The study team shared these data at monthly meetings with all DSD staff at the facility to review progress, identify challenges, and to facilitate the use of quality improvement methods to develop and test adjustments as needed. Notes from these meetings and discussions were used to synthesize key lessons learned, we classified the success as enabling factors and challenges as hindering factors of implementing this approach. The implementers, facility-based clinicians and community health workers assessed the tools and processes to be effective and relatively efficient.

### Data Collection and Management

All participants were monitored for possible IPT related side effects, phone and in-person appointment keeping, and IPT adherence and completion. Existing data collection systems and tools such as the electronic medical records (EMR) and ART registers were adapted to capture and record project data from month 3-9 of implementation (September 2019-April 2020) (Figure 2). In May 2020, pilot staff trained in research ethics and participant confidentiality extracted participant demographics, reported IPT-related side effects, appointment keeping and IPT completion from the EMR. Data was checked for completeness, cleaned, and stored in a secured and password-protected PostgreSQL database for analysis. We de-identified the data and only included fields necessary for analysis.

### Data Analysis

The primary outcome was self-reported IPT completion defined as completing the dispensed 6-month course of IPT. Secondary outcomes were reported side effects likely associated with IPT (burning sensation in fingers and toes; itchy skin, yellow eyes, tongue and palms; joint pains; nausea; vomiting; stomach pains and fever) and the percentage of scheduled phone appointments completed. We used descriptive statistics including frequencies and proportions to describe participant characteristics such as age, gender, time on ART and outcome variables. Fisher’s exact test was used to determine if participant demographics were statistically associated with primary and secondary outcomes.

### Ethical Consideration

The University of Zambia Biomedical Research Ethics Committee (UNZABREC) approved, and the National Health Research Authority (NHRA) gave authority to conduct the pilot. Since this evaluation was implemented within routine ART program, signed informed consent was not a requirement for participation.

## Results

### Participant Characteristics

A total of 1167 PLHIV were screened, of whom 818 (70.1%) were eligible and enrolled within the 30-day period. Of the 349 (29.9%) excluded, 172 (14.7%) had no phone; 122 (10.5%) were already on IPT; 30 (2.6%) had recently completed IPT/TB treatment; 13 (1.1%) had TB symptoms; 7 (0.6%) were pregnant; and 5 (0.4%) declined to participate in the pilot (Figure 4). The 818 participants had median age of 42 years (IQR=35, 49), and 540 (66%) were females. Over half (56.6%) of the participants were on ART for at least 5 years (Table 1).

**Figure 4:**
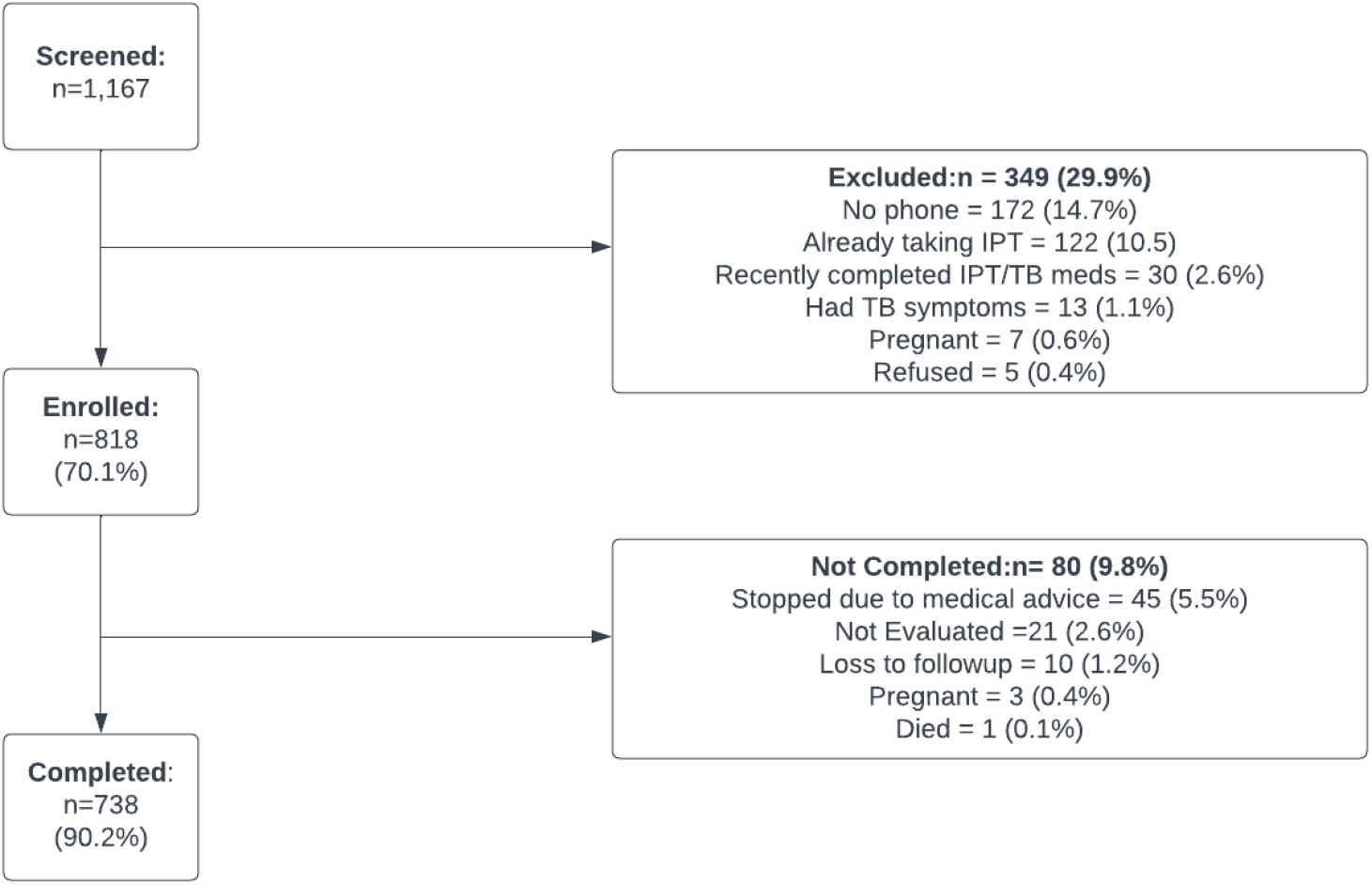
Enrollment Flow Chart.

**Table 1:** Baseline characteristics and outcomes of enrolled participants.

| Characteristics | Number of participants (% of Total) | Number (%) completed treatment | P-value | Number (%) Kept All Appointments | P-value | Number (%) Experienced Side Effects | p-value |
| --- | --- | --- | --- | --- | --- | --- | --- |
| **Age (years): median (IQR)** | 42, (35-49) | 42 (35-49) |  | 42 (35-49) |  | 44.5 (38-49) |  |
| <30 | 89 (10.9%) | 77 (88.5%) | 0.22 | 60 (67.4%) | 0.91 | 2 (2.3%) | 0.2 |
| 30-39 | 270 (33%) | 242 (89.6%) |  | 174 (64.4%) |  | 18(6.7%) |  |
| 40-49 | 286 (35%) | 263 (91.0%) |  | 192 (67.1%) |  | 30 (10.5%) |  |
| 50+ | 173 (21.15%) | 156 (90.2%) |  | 113(65.3%) |  | 16 (9.25%%) |  |
| **Sex** |  |  | 0.73 |  | 0.81 |  | 0.5 |
| Female | 540 (66%) | 484 (89.6%) |  | 354 (65.6%) |  | 167(31.0%) |  |
| Male | 278 (34%) | 254 (91.4%) |  | 185 (66.6%) |  | 86 (31.0%) |  |
| **ART Duration (years)** |  |  | 0.20 |  | 0.5 |  | 0.5 |
| 1-2 | 61 (7.5%) | 52 (85.3%) |  | 37 (60.1%) |  | 3(5%) |  |
| 3-5 | 153 (18.7%) | 139 (90.9%) |  | 106 (69.3%) |  | 10(6.6%) |  |
| 5+ years | 463 (56.6%) | 418 (90.3%) |  | 299 (64.6%) |  | 37 (8%) |  |
| Missing | 141 (17.2%) | 129 (91.5%) |  | 97 (68.8%) |  |  |  |
| **Total** | **818 (100%)** | **738 (90.2%)** |  | **539 (65.9%)** |  | **66(8.1%)** |  |

### IPT completion

Of the 818 enrolled participants, 738 (90.2%) reported completing the full 6-month course of IPT; there were no significant differences in completion rate by age, sex, or duration on ART (Table 1). Of the 80 (9.8%) of participants who did not complete the full course of IPT, 45 (5.5%) initiated IPT and then stopped due to medical advice as they experienced side effects, 21 (2.6%) were not evaluated,10 (1.2%) were lost to follow-up and 3(0.4%) became pregnant (Figure 4). One participant died in week 1 of the study but had not initiated IPT according to their family.

### Keeping scheduled appointments

Five hundred and thirty-nine (65.9%) participants kept all seven phone appointments; there were no significant differences in completion rate by age, sex, or duration on ART (Table 1). Appointment keeping was lower in the first three months of IPT initiation. (Figure 5). Approximately 20% of participants did not keep their phone appointments at weeks 2, 4 and 8. Following two failed phone calls on two separate days a physical visit was then conducted, on average, 75% of these were reached physically and of these 95% reported adherence with IPT.

**Figure 5:**
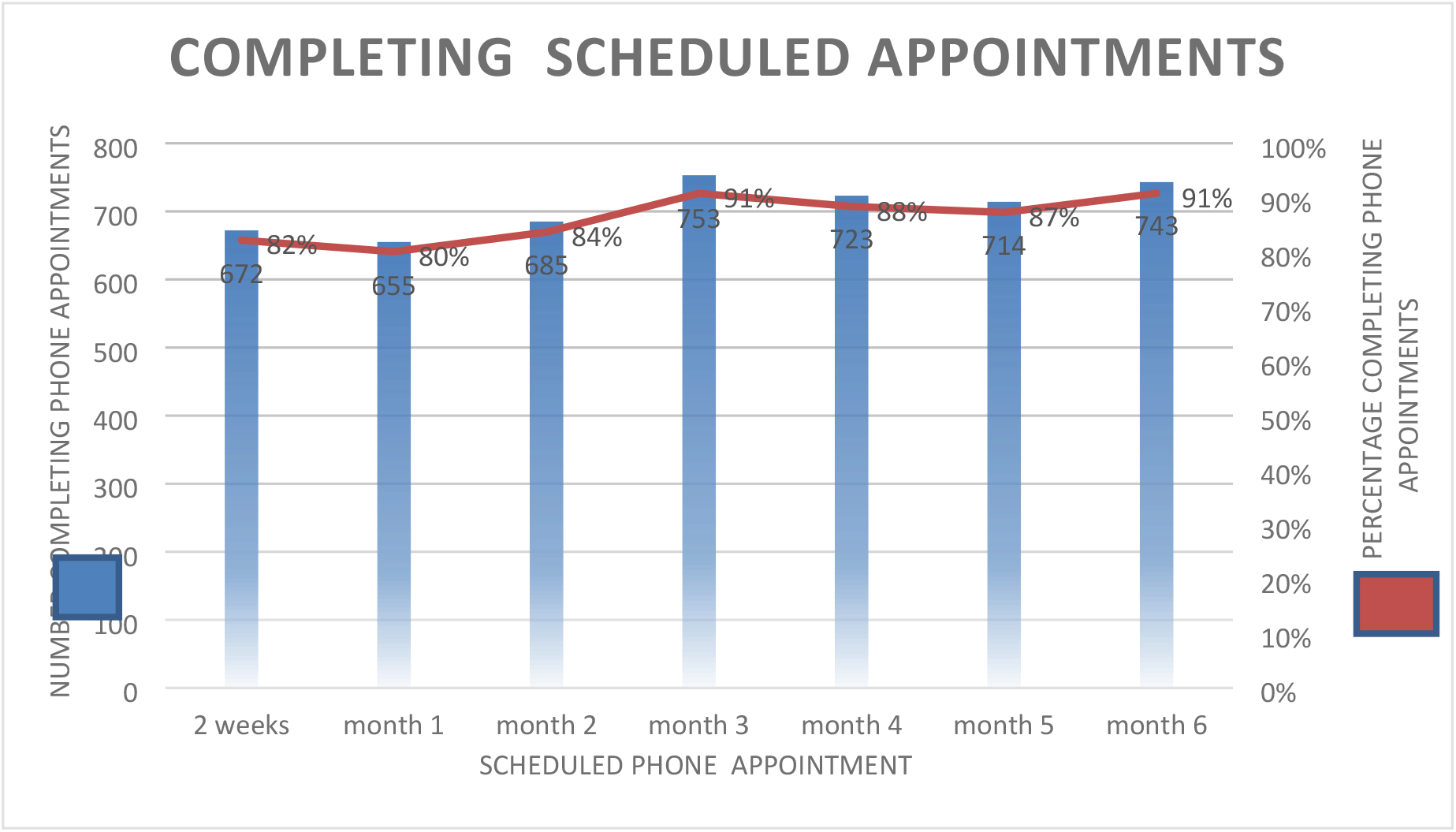
Completion of Phone Visits by Month.

### Reported Side Effects

Sixty-six participants (8.1%) reported side effects; there were no significant differences by age, sex, or duration on ART (Table 1). Reports of side effects were more frequent in the early weeks of IPT (Figure 6). Whilst none of the side effects were classified as serious adverse event according to Zambia Medicines Regulatory Authority (ZAMRA) definition,^22^ forty-five (5.5%) of participants were advised by their healthcare workers to discontinue IPT for fear of side effects worsening during IPT administration. (Figure 4).

**Figure 6:**
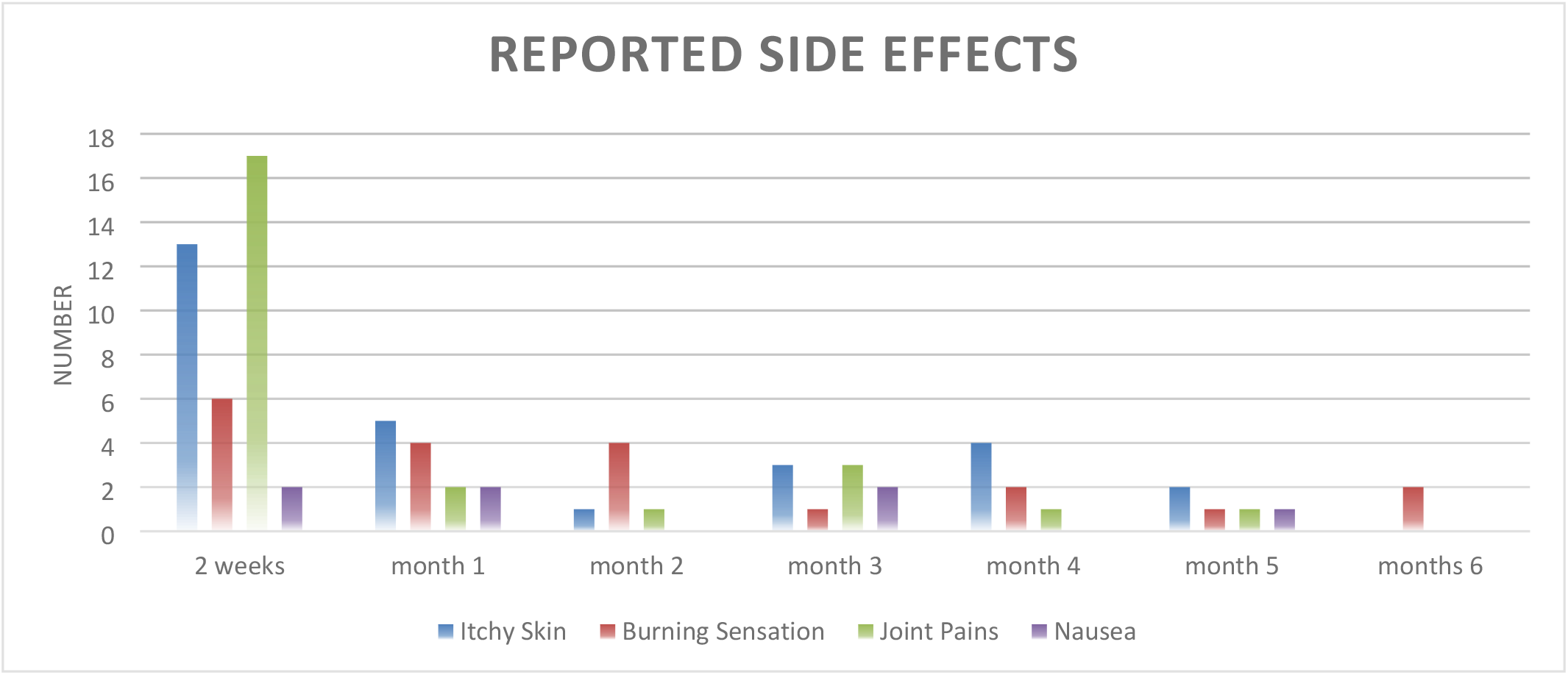
Reported Side Effects.

### Lessons learned: Successes

#### Effectively adapting existing processes and clinical tools

Adapting existing process and clinical tools ensured identification of the appropriate facility staff to provide screening, counselling, enrolment, IPT initiation and follow-up services. Additionally, it harmonised the tools (screening tools, clinical records and SOPs) thus minimized burden on facility staff and ensured that the intervention was not viewed as an additional responsibility.

#### Demand Creation and Client Empowerment

CHWs reported that the strategy of utilizing structured (1) educational messages messages and (2) checklist and script empowered them to generate demand for the intervention, to allay participants’ concerns and consequently empowered participants to monitor and take charge of their health.

#### Cultivating a collaborative structured learning environment

The monthly meetings at the facility cultivated an iterative and collaborative learning environment that promoted and provided a platform for initial and ongoing success of integrating IPT into FT models.

#### Promoting ownership

The HCWs showed absolute involvement and support for this pilot project, which enabled the implementation of this new approach.

##### Ensuring commodity security

Securing the supply chain system ensured constant availability of both INH and Pyridoxine (Vitamin B6).

### Lessons learned: Challenge

#### Phone usage

The intervention model included 7 phone check-ins over the 6-month intervention, meaning that access to phones was critical for pilot implementation and effectiveness. Although access to a phone was an eligibility criterion, approximately 20% of participants did not keep their appointments in week 2, months 1 and 2 as phones were either switched off or went unanswered despite repeated attempts. For those not reached by phone, CHWs conducted in-person follow ups to ascertain IPT adherence, about 75% were reached physically of whom 95% reported taking IPT as prescribed. During the in-person follow-ups, CHWs emphasized the importance of the phone follow-ups this improved phone usage in subsequent months (Figure 5).

## Discussion

This pilot represents one of the first examples of integrating long refills of TPT (6MMSD) into a facility-based DSD model with utilization of phone calls at shorter intervals for continuous education, close monitoring and adherence support. We found that integration of TPT uptake amongst participants was considerably higher than the national average, and self-reported TPT completion rates were similar to those found nationwide. Our project builds on examples from case studies findings conducted in South Africa and the Democratic Republic of Congo that indicated integration of TPT into DSD models could be successfully adopted as a TB screening strategy and that longer refills of TPT were feasible.^14,21^ Additionally, findings of high IPT completion rates and low reports of IPT side effects from this pilot are consistent with early findings from the implementation of IPT in DSD models in rural Uganda.^24^

In this pilot project we found that 90.2% of the participants who accepted this innovation completed their IPT, similar to Zambia’s reported national completion rate of (90%).^25^ Although design of this study did not include control or comparison groups, this high completion rate is encouraging and may have been due to the combination of program efficiency and access to ongoing phone-based counselling and support, which we believe is an innovative strategy of facilitating social support. Studies by Grimsrud *et al*. and Nachenga *et al*. underscore the significant contribution of social support in promoting IPT completion in community-based HIV DSD models. ^26,27^ Further observations made by authors from South Africa and Uganda, where higher ITP completion rates were reported in clients exposed to enhanced social support in both DSD community-based models and standard of care. ^21,24, 28^ Whilst our pilot integrated IPT in a facility-based model, we believe that the continuous social support provided via phone promoted treatment completion.

Furthermore, high completion rates seen in our pilot are concordant with evidence from a quality improvement collaborative in Uganda that showed 89% completion rates and two studies conducted in Swaziland and Uganda that showed improvement of TPT completion in models that integrated TB and HIV services.^21,28,30^ In Swaziland, IPT completion rates in facility based (FB) models and community based (CB) models was reported at 89% and 91% respectively.

Both models utilized trained HCWs to provide support and counselling. ^28^ Similarly, the Ugandan cross-sectional study showed a higher completion rate in people in DSD models (72%) than in routine care (53%). Results from this study suggested that the DSD models allowed for stronger HCW-client relationship, education, communication and empowerment. ^21^

Uptake and coverage of IPT amongst PLHIV in Zambia ranges from 18% to 49% while in this pilot 70.1% of all enrolled participants accepted the intervention.^4,31^ We hypothesize that the person-centered and convenient model being piloted led to higher than usual 70.1% uptake rate. DSD models that extended clinic visits have shown similar uptake success in HIV programs ^32-34^ thus, supporting clients preference for extended integrated visits.

Given the push to leverage DSD models to scale up TPT for PLHIV, it was important to evaluate TPT in routine care, particularly in relation to side effects and treatment completion.^37^ Collecting and evaluating side effects data will capacitate the HCWs to reasonably inform patients regarding the benefits versus risks of TPT. ^35^HCWs have often conveyed fears of IPT medication toxicity due to concomitant use with ART. ^36,37^ Our evaluation data showed that at 6 months of IPT initiation only about 8.1% of the participants experienced side effects thus suggesting the fears to be unfounded. These findings are corroborated by the 2019 Centre for Global Health Division TPT myth and fact, which concluded that risk of adverse events in patients on TPT was low, with <10% side effects recorded from various studies.^37^ Additionally the national QI collaborative in Uganda similarly reported that <10% of the study participants experienced INH adverse events.^30^

This proof-of-concept pilot shows the importance of utilizing existing processes, establishing HCWs project ownership, promoting client empowerment and fostering iterative and collaborative learning environment all of which were pivotal to the success. A 2019 debate of lessons learnt during IPT implementation in Zambia closely linked programmatic issues such as HCWs capacity to manage patients, limited demand for TPT services, drug stock outs and patient IPT knowledge to the IPT completion and scale up.^22^ A week supply chain is a significant hinderance to treatment completion and TPT scale up.^26,27^ In the prospective cohort of improving IPT delivery in DSD models in Swaziland, erratic IPT supply was one of the reasons for non-completion of TPT.^29^ Therefore, securing the supply chain was useful for implementation and maintenance of this pilot.

Strengths of this program evaluation include the innovative pilot design that does not only reduce the burden of patients frequenting the health facility, but moreover promotes MoH recommendation of minimizing PLHIV side effects risks by ensuring exhaustive education of the patients and close monitoring. ^2^Additionally provision of IPT long refills and utilization of phone calls in between clinic visits is an intervention we believe may be leveraged to support uptake and completion of other TPT regimens such as Isoniazid plus Rifampicin (3HP) and Isoniazid plus Rifampicin (3HR) in Zambia.^2^ Another strength of this pilot was the documentation of the implementation process. Limitations include those typically associated with a single-site proof-of-concept project of this kind: the absence of a control or comparison group other than historical experience with IPT in Zambia, lack of TB infection outcomes reliance on self-report to assess IPT completion, and the purposive selection of one urban health facility and the exclusion of PLHIV without phones, all of which limit the strength and generalizability of our results. The description of lessons learnt draw on the experiences of the project team, and while we believe it is important to share these experiences, we also recognize the subjective nature of the successes and challenges reported. Nonetheless, we are confident that the results of this pilot project will contribute to the growing body of literature on the importance of integrating TPT and other health services into DSD models for people with HIV.^38^

## Conclusions

This pilot demonstrated that an integrated MMSD/IPT model involving synchronized IPT and HIV clinical visits, dispensing 6 months of IPT and ART, and delivering adherence counseling and follow up via phone was associated with excellent IPT uptake and completion rates. This promising public health strategy is worth further study and may improve coverage and completion of TPT amongst PLHIV, especially in countries that have taken DSD to scale and in which mobile phone penetration is high. Fostering project ownership, empowering clients, building health care worker capacity, collaboration with key stakeholders, mobilizing human resources and attending to supply chain security, were essential to successful pilot implementation.

## Data Availability

The Government of Zambia allows data sharing when applicable local conditions are satisfied. In this case, the data from the study will be made available to any interested researchers upon request. The CIDRZ Ethics and Compliance Committee is responsible for approving such request. To request data access, one must write to the Committee chair/Chief Scientific Officer, Dr. Roma Chilengi, or the Secretary to the Committee/Head of Research Operations, Ms. Hope Mwanyungwi mentioning the intended use for the data. The Committee will then facilitate review and authorization to release the data as requested. Data requests must include contact information, a research project title, and a description of the intended use.

## Authors’ contributions

MMM: Lead author, project design, data analysis and wrote the first draft

MM: Author, field coordination and data collection

PS: Conducted data analysis

KZ: Assisted with project conceptualization, project design and manuscript writing

MS: Assisted with and project design

NZ: Assisted with and project design

EK: Assisted with data collection and pilot implementation

LKM: Assisted with project conceptualization and design

MR: Assisted with manuscript development and writing

FM: Assisted with project conceptualization and design

FM: Assisted with project conceptualization and design

FC: Assisted with project conceptualization and design

FM: Assisted with project conceptualization, design and implementation

PP: Assisted with project conceptualization and design

CBM: Advised with the implementation process

SB: Lead advisor on program evaluation and data analysis

AS: Assisted with manuscript writing

KM: Assisted with project conceptualization, design and implementation

PK: Pilot conceptualization

LM: Technical guidance and pilot conceptualization

PL: Technical guidance and pilot conceptualization

PM: Assisted with project conceptualization, design and writing

## Acknowledgements

We are indebted to the Government of the Republic of Zambia, through the Ministry of Health, for their valuable guidance and support during implementation. We would like to thank the DSD Task Force and the TB National Program for leading the conceptualization and providing meaningful guidance throughout. We are grateful to ICAP at Columbia University and the CQUIN Learning Network for this great initiative that culminated into novel findings. Lastly, we thank and salute the participants and health care workers for their efforts and contributions to the larger vision of achieving epidemic control.

## Copyright

This is an open access article distributed under the terms of the <u>Creative Commons Attribution License</u>, which permits unrestricted use, distribution, and reproduction in any medium, provided the original author and source are credited.

